# Deriving Optimal Treatment Timing for Adaptive Therapy: Matching the Model to the Tumor Dynamics

**DOI:** 10.1101/2025.04.01.25325056

**Authors:** Kit Gallagher, Maximilian A. R. Strobl, Alexander R. A. Anderson, Philip K. Maini

## Abstract

Adaptive therapy (AT) protocols have been introduced to combat drug-resistance in cancer, and are characterized by breaks in maximum tolerated dose treatment (the current standard of care in most clinical settings). These breaks are scheduled to maintain tolerably high levels of tumor burden, employing competitive suppression of treatment-resistant sub-populations by treatment-sensitive sub-populations. AT has been integrated into several ongoing or planned clinical trials, including treatment of metastatic castrate-resistant prostate cancer, ovarian cancer, and BRAF-mutant melanoma, with initial clinical results suggesting that it can offer significant extensions in the time to progression over the standard of care.

However, these clinical protocols may be sub-optimal, as they fail to account for variation in tumor dynamics between patients, and result in significant heterogeneity in patient outcomes. Mathematical modeling and analysis have been proposed to optimize adaptive protocols, but they do not account for clinical restrictions, most notably the discrete time intervals between the clinical appointments where a patient’s tumor burden is measured and their treatment schedule is re-evaluated. We present a general framework for deriving optimal treatment protocols which account for these discrete time intervals, and derive optimal schedules for a number of models to avoid model-specific personalization. We identify a trade-off between the frequency of patient monitoring and the time to progression attainable, and propose an AT protocol based on a single treatment threshold. Finally, we identify a subset of patients with qualitatively different dynamics that instead require a novel AT protocol based on a threshold that changes over the course of treatment.

## 1 Introduction

Adaptive therapy (AT) is a new cancer treatment paradigm that uses principles from evolution to delay disease progression for late-stage cancer patients. Cancerous tumors are highly genetically heterogeneous (Allison and Sledge, 2014), comprised of many different clones with potentially differing resistance to any given therapeutic agent (Turner and Reis-Filho, 2012). This heterogeneity enables tumors to be considered through the lens of Darwinian evolution, wherein multiple species with different (environment-dependent) fitness compete with each other (Gerlinger and Swanton, 2010). In particular, competition (be it for resources, space to grow, or access to vasculature) between clones of variable drug resistance can reduce growth rates, and result in ‘competitive suppression’ of all clones, including drug-resistant clones that are otherwise unencumbered by the application of treatment. Suppression may be further enhanced by naturally lower base proliferation rates of drug-resistant cells relative to sensitive cells (a ‘cost of resistance’), as observed *in vivo* across several tumor cell lines (Enriquez-Navas et al., 2016; Bacevic et al., 2017).

Treatment breaks/holidays may leverage this competitive suppression to gain indirect control over drug-resistant cells. While drug-sensitive clones can be controlled directly by the application of treatment, the growth of resistant clones can only be limited indirectly, via competitive suppression from the drug-sensitive cells. These treatment breaks allow regrowth of the drug-sensitive population, renewing the competitive suppression of resistant cells and re-sensitizing the tumor to treatment. In contrast to previous clinical implementations of intermittent therapy (Hussain et al., 2013; Crook et al., 2012), wherein routine drug holidays of a fixed length were scheduled uniformly across the whole patient cohort, AT tailors drug scheduling to an individual patient’s tumor dynamics (Gatenby et al., 2009). These adaptive strategies exploit both spatial and resource competition between drug-sensitive and -resistant cells (Gallaher et al., 2018; Bacevic et al., 2017; Strobl et al., 2022).

A comparison between continuous treatment (CT) and adaptive therapy is given in Figure 1a - the conventional AT protocol applies treatment until the tumor halves in size, before withdrawing treatment until the tumor regains its initial size. We will refer to this formulation of AT as ‘window-based’ AT, as the tumor size is maintained within a set window based on the initial tumor size. This protocol is termed AT50, as treatment is applied until the tumor size is 50% of the original size, and these treatment cycles are repeated until the tumor grows 20% larger than the initial tumor size, when it is deemed to have progressed. The ‘window-based’ AT protocol has been applied clinically to metastatic, castrate-resistant prostate cancer, where a two-fold increase in the mean time to progression (TTP) was observed, broadly in agreement with the simulated benefit here (Zhang et al., 2022). An example of the patient dynamics from this trial is given in Figure 1b. The tumor size is tracked via prostate-specific antigen (PSA) levels, a common proxy for tumor burden in prostate cancer.

**Figure 1:**
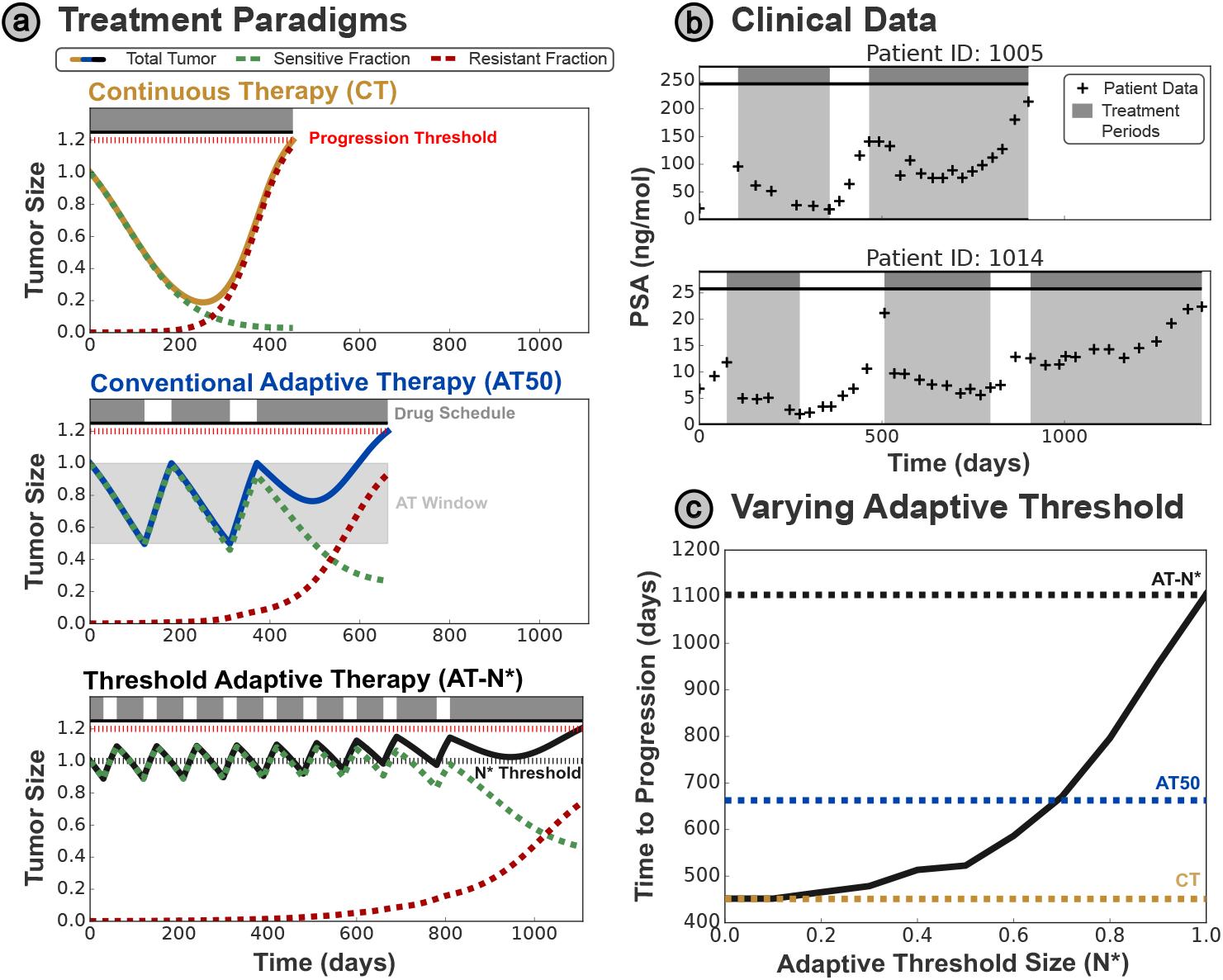
**(a)** Comparison of different treatment protocols - in each case tumor progression is defined as 20% growth from the initial size. The tumor size has been normalized relative to the initial tumor size in each plot. The clinical standard of care is based on continuous treatment, however, progression can be delayed by introducing breaks in treatment - in the conventional (‘window-based’) AT a treatment holiday is initiated when the tumor has halved in size, and resumed when the tumor returns to the initial size. We propose a modified AT protocol (AT-N*) based on a single threshold *N*^*^ - treatment is only delivered when the tumor is larger than this threshold. Overshooting of the threshold occurs as treatment may only be modified every 30 days. **(b)** We illustrate the clinical application of AT, showing the outcomes on two exemplar patients from a clinical trial on metastatic prostate cancer based on the AT50 protocol (Zhang et al., 2022). PSA is used as a clinical biomarker for the total tumor burden in this disease setting. **(c)** Higher treatment thresholds correspond to greatly delayed progression, with significant improvement over the standard of care.

We contrast these ‘window based’ approaches with our proposed ‘threshold-based’ strategy (AT-N*), wherein treatment is only given when the tumor is larger than a pre-specified threshold size (*N*^*^). In Figure 1a the initial tumor size is used, resulting in a significant further increase in the TTP relative to CT and AT50.

These benefits are achieved by maintaining a larger sensitive cell population to continually suppress the growth of resistant cells. This is illustrated in Figure 1c — by varying the threshold size *N*^*^ used for AT-N*, we see that higher thresholds that maintain a larger sensitive cell population result in greater TTP values. This is supported by previous analytic work, which shows that the TTP may be maximized by maintaining as large a sensitive sub-population as possible (Hansen et al., 2017; Viossat and Noble, 2021), below some upper bound on the tumor size (given by the progression limit). Hansen and Read have even proposed setting the upper limit of ‘window-based’ AT above the initial size (Hansen and Read, 2020), based on the proposal that patients may tolerate slight increases in the tumor burden above the initial size, and this has since been implemented as ‘Range-Bounded’ Adaptive Therapy (Brady-Nicholls and Enderling, 2022).

However, these studies assume continuous monitoring of the tumor size, allowing the drug to be withdrawn/reapplied as soon as the tumor size crosses a pre-defined threshold. With the development of wearable devices this may be a realistic assumption in the future, however it is not currently possible in the clinic. We show in Section 2.5 that these optimal frameworks break down in clinically realistic settings, where updates to the treatment schedule can only be made at discrete time points, and the tumor size is not tracked between these time points. These limitations motivate revision of previous optimal AT protocols, which relied upon continuous monitoring and decision-making capability to maintain a maximal sensitive subpopulation.

In this paper, we present a framework to derive an optimal treatment threshold based on the discrete time interval between treatment updates, before applying this framework to a range of mathematical tumor models with different underlying assumptions. In each model we consider, the cancer is incurable since there is a drug-resistant species that cannot be fully eliminated, and the null state (*S* = *R* = 0) is unstable. This is a common phenomenon in metastatic cancers with existing treatment-resistance; instead of seeking to cure the cancer, we therefore aim to control its growth to increase the TTP. The models we discuss below were all previously proposed to describe adaptive therapy in metastatic prostate cancer.

We find that accounting for discrete appointment intervals requires highly patient-specific optimal thresholds, motivating the need for personalized approaches to adaptive therapy. This paper provides an analytic approach to evaluating patient-specific optimal treatment protocols based on the parameters of the tumor dynamics. We also show that the optimal treatment protocols depend on the underlying model assumptions — for example a model that captures systematic changes in the patient’s tumor dynamics over time requires a time-varying optimal treatment threshold. Overall, we develop an approach to incorporate clinical realities into mathematical analysis of optimal treatment schedules, and present optimal treatment approaches for a range of competition-based tumor models.

## 2 Methods

In this section, we introduce three distinct ordinary differential equation models for the response of a heterogeneous tumor to therapy. In each case, the heterogeneity in drug response is captured by two separate populations, which are subject to some form of inter-species competition. Each model was previously developed to capture the drug response dynamics of patients undergoing intermittent/adaptive scheduling of hormone therapy for metastatic prostate cancer.

### 2.1 Lotka–Volterra Model

Strobl et al. (2021) model heterogeneity in drug response within a partially drug-resistant tumor by two competing cell types: drug-sensitive cells *S*(*t*), and fully resistant cells *R*(*t*), via the following two population Lotka–Volterra model (Lotka, 1910; Volterra, 1928):

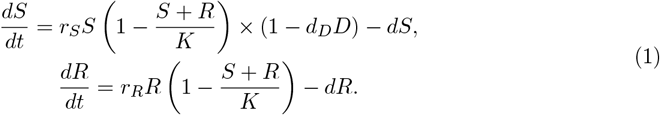

Competition is represented by a logistic growth model with a shared carrying capacity *K*, while each species has a separate growth rate (*r*_*S*_ and *r*_*R*_ respectively). Cells are assumed to die naturally at the same rate *d*, while the Norton–Simon model (Norton et al., 1977) is adopted to model the drug-induced killing of sensitive cells, occurring at a rate proportional to the population’s growth rate and the drug concentration, *D*(*t*) (with proportionality factor *d*_*D*_).

This model only considers pre-existing resistance, and there is no mechanism for the acquisition of drug resistance during treatment. However, the inclusion of genetic mutations in simple models has been shown to have no significant bearing on the overall response to therapy (Viossat and Noble, 2021). Parameter values were adopted from Strobl et al. (2021) and are given in Table S1. Note that throughout this paper, we will use *N* to denote the total tumor size - i.e. *N* = *S* + *R*.

### 2.2 Waning Competition Model

In comparison, we also consider a modified Lotka–Volterra model proposed by Lu et al. (2024). This time-varied generalized Lotka–Volterra model (with growth scaling exponent *α*) also considers separate drug-sensitive and drug-resistant cell populations competing for shared resources in the tumor microenvironment. However, Lu et al. (2024) model an exponentially decreasing trend in resource overlap, with the resistance index *γ >* 0 (hence this model will be referred to hereafter as the ‘Waning Competition’ model). They attribute this trend to competition-induced mutations and epigenetic modifications within the cancer population, such that competition intensity weakens over time as there are fewer shared resources. Explicitly, this model may be written (in non-dimensional form) as:

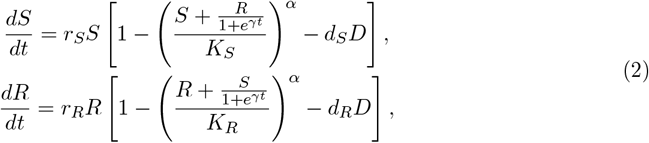

where *S* and *R* are the sensitive and resistant cell sub-populations respectively, and *D* is the drug concentration. Each cell species *i* has a distinct growth rate *r*_*i*_, carrying capacity *K*_*i*_ and drug-induced death rate *d*_*i*_, while logistic growth accounts for net growth. It is worth noting that this model also reduces to the generalized logistic model in the case where *γ* = 0 (eliminating the explicitly time-dependent competition), and in the absence of treatment.

For this model, progression was defined as growth in the resistant population alone to 0.1*K*_*R*_, replicating the definition used by Lu et al. (2024). The model was parameterized using values from Lu et al. (2024) (specified in Table S2).

### 2.3 Stem-Cell Model

In contrast to the previous models, which categorize cells by their drug response, we also considered an alternate model proposed by Brady-Nicholls and Enderling (2022), which distinguishes between prostate cancer stem-like (*S*) and differentiated (*D*) cells to model the tumor response to treatment. Stem-like cells divide at rate *λ*, to produce either two stem-like cells (with probability *p*_*s*_, but subject to negative feedback 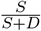 from differentiated cells), or a stem-like and a non-stem cell. While stem-like cells are androgen-independent and hence do not respond to treatment, differentiated cells die in response to drug application at rate *d*_*D*_. A binary dosing schedule is implemented through *T*_*x*_, where *T*_*x*_ = 1 corresponds to treatment being given, and *T*_*x*_ = 0 for treatment holidays.

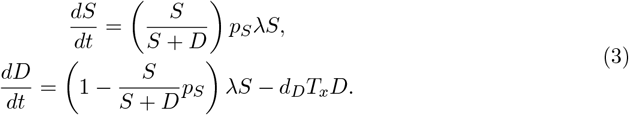

The model was parameterized using values from Brady-Nicholls and Enderling (2022) (specified in Table S3).

### 2.4 Adaptive Therapy

We consider three distinct treatment protocols for the drug concentration *D*(*t*) (or *T*_*x*_(*t*) for the Stem-Cell model) within this work. All drug doses are normalized such that *D*(*t*) = 1 corresponds to the maximum tolerated dose.

1. **Continuous Therapy (CT)** – The standard of care:

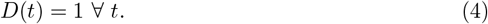
2. **Window-Based AT (AT50)** – Treatment is given until a decrease to 50% of the initial size (*N*_0_) is achieved, then withdrawn until the tumor returns to its initial size. AT50 was used in the pilot AT clinical trial by Zhang et al. (2017, 2022):

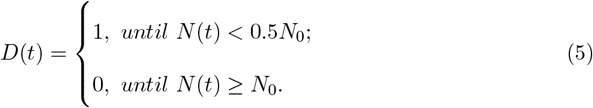
3. **Threshold-based AT (AT-N*)** – Treatment is given only when the tumor is larger than a set threshold size *N*^*^:

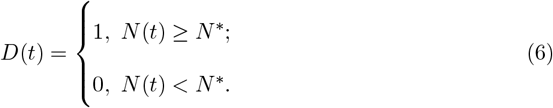

Treatment outcomes from these schedules are compared according to their TTP, where progression is defined as a 20 % growth from the initial size (i.e. 1.2*N*_0_), as in prior studies in this area (e.g., Gallaher et al. (2018); Strobl et al. (2021); Viossat and Noble (2021)).

### 2.5 Optimal Threshold

We now consider the case of clinically realistic treatment protocols, limited by discrete time monitoring, and introduce the notion of an optimal threshold. The optimal threshold may be derived by considering a phenomenon we define as ‘premature progression’ - where insufficient treatment (or an overly long treatment holiday due to discrete time monitoring) results in progression due to growth in the sensitive cell population. This is deemed ‘premature’ as the tumor is still (partially) sensitive to treatment, and so progression could have been delayed by more frequent re-evaluation of treatment. Figure 2a demonstrates that even small increases in the interval between appointments may transform a successful strategy into one that undergoes progression in the first treatment cycle, with the tumor size *N* (*t*) jumping from *N* (*t*) *<* 0.5*N*_0_ (the threshold for treatment in conventional AT50) to *N* (*t*) *>* 1.2*N*_0_ (the threshold for progression) in a single treatment cycle.

**Figure 2:**
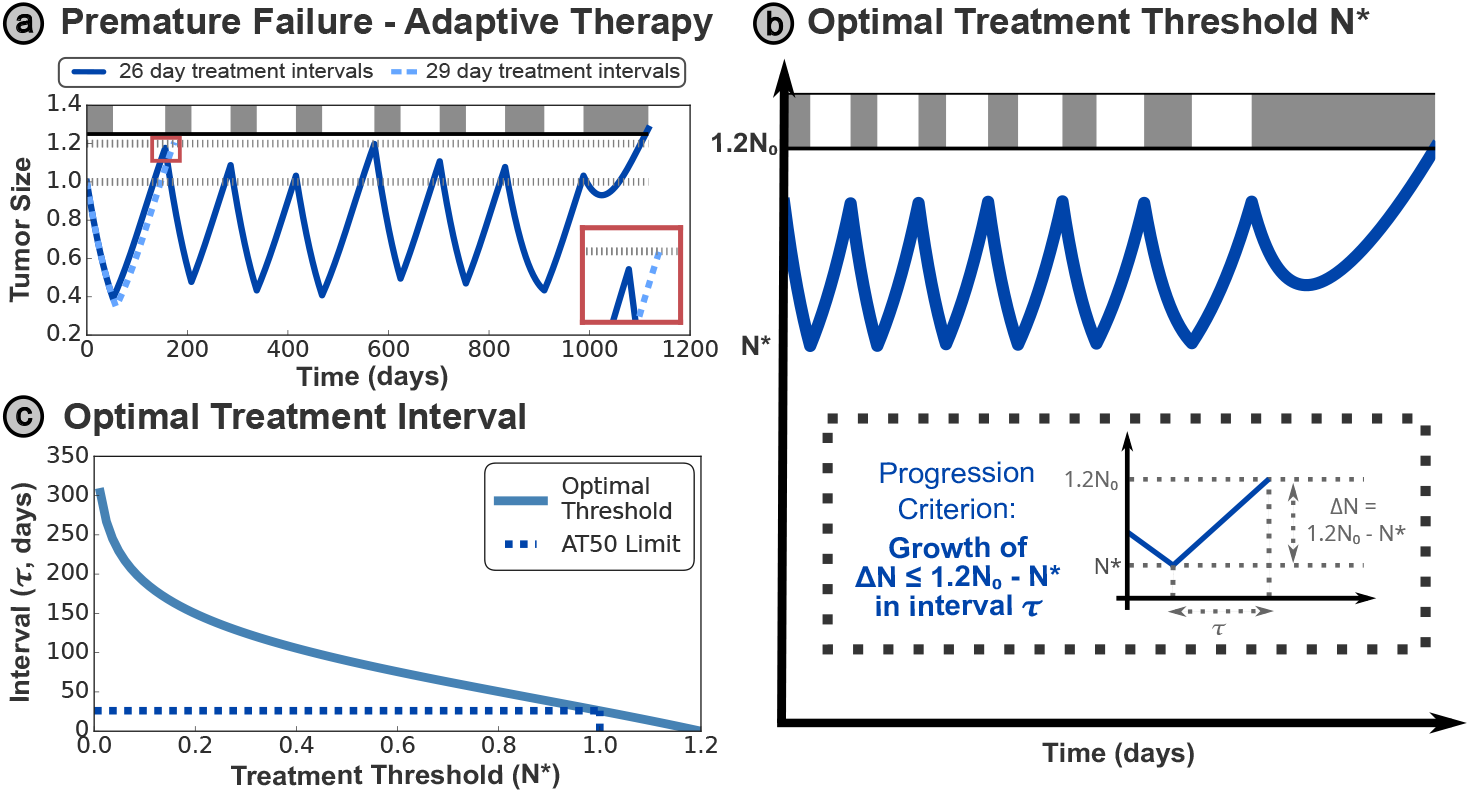
**(a)** Premature progression occurs when treatment decisions are made insufficiently frequently, such that the sensitive fraction of the tumor can grow from below the treatment threshold (*N* (*t*) *<* 0.5*N*_0_) to above the progression threshold (*N* (*t*) *>* 1.2*N*_0_) within a single treatment cycle. This places an upper limit on the maximum treatment threshold, given patients are monitored at discrete intervals in the clinic. In this case, a small increase in the appointment interval (from 26 to 29 days) resulted in premature progression - shown in detail in the red inset - which causes a significant decrease in TTP. This figure uses a set of modified parameter values for visualization purposes, with *N*_0_ = 0.3, *d*_*S*_ = 0.3*r*_*S*_. **(b)** We can derive the optimal treatment threshold *N*^*^ for a particular appointment interval *τ* by ensuring that the time taken for the tumor to grow from the threshold to the progression limit is more than the time interval between appointments (see text for details). **(c)** The required interval decreases monotonically as the treatment threshold is increased, however, this relationship is non-linear. The upper limit threshold of *N*_0_ from conventional (‘window-based’) AT50 is indicated by the dashed line — this corresponds to an analytic limit on the AT appointment interval of 26.1 days, supporting the observation in (a).

We define the critical treatment time (*τ* in Figure 2b) as the maximal possible time interval between treatment appointments, to avoid premature progression at a given treatment threshold. This may be obtained as the time for the tumor to grow from the threshold size for treatment to the progression limit. While the details of evaluating this differ between models, and often require some simplifying assumptions such as a negligible resistant population, the overarching approach is common between all models.

An example of this growth time for the Lotka–Volterra model (derived in Supplementary Section S3.1) is plotted in Figure 2c - the optimal threshold decreases monotonically as the appointment interval increases, though this relationship is not linear. In practice, the interval between appointments is often determined by clinical availability and practical restrictions, so it is more practical to personalize the threshold tumor size *N*^*^ within the AT protocol to each patient, based on a fixed time interval *τ*.

We will apply this approach to each of the three tumor models introduced above, illustrating its applicability to different modeling frameworks. While the mathematical forms of the optimal threshold differ between models, we can identify key qualitative trends in the clinical data which inspired these models.

## 3 Results

### 3.1 Lotka–Volterra Model

The aim of this paper is to develop tools to improve patient outcomes by personalizing their treatment schedules. Before detailing how we propose to optimize scheduling, it is important to discuss the optimality criterion by which we seek to improve outcomes. Given the presence of a fully-drug resistant population, it is not possible to eliminate the tumor entirely, which would cure the cancer. Furthermore, we show in Supplementary Section S2 that all the non-zero stationary states of the tumor are on the order of the carrying capacity *K*. We treat this size as an intolerable burden, where the tumor has taken over a host completely and no more resources are available for growth. For this reason, in Section 2.4 we restrict the total tumor size *S*(*t*) + *R*(*t*) = *N* (*t*) *<* 1.2*N*_0_, where *N* (*t*) = 1.2*N*_0_ corresponds to clinical progression. It is inevitable that the tumor will ultimately reach this progression limit, and hence we also cannot contain the tumor at a tolerable size indefinitely. An optimal treatment strategy should therefore aim to delay the tumor growth, such that the time taken to reach this progression limit is maximized.

The net tumor growth rate is minimized when *N* is large, and so we adopt a threshold-based AT protocol, as further motivated in Supplementary Section S2.4. This is based on a threshold *N*^*^, as introduced in Section 2.5. By computing the maximal possible growth in a single appointment interval *τ*, as described in Section 2.5, we derive the following expression for *N*^*^ (full derivation given in Supplementary Section S3.1):

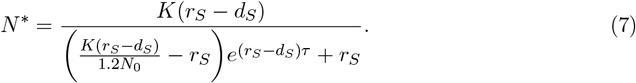

Plotting this relationship in Figure 3a, we can see that the higher treatment thresholds necessitate more frequent appointments in general, and the analytically optimal threshold of 1.2*N*_0_ is only possible when treatment is being continuously re-evaluated (*τ* = 0). For contrast, an AT50 approach would only be possible if treatment is re-evaluated at least every 40 days, and longer intervals would risk premature progression. In the inset panels, we contrast the optimal treatment schedules for monthly (30 day) and bimonthly (60 day) appointment intervals. The shorter interval allows better control over the tumor size, maintaining a more consistent, and higher average, size than the longer interval. The increased average tumor size results in greater suppression of the resistant cells, and hence greater overall TTP, but require more frequent appointments to maintain. In other words, there is a trade-off between the cost and inconvenience of more frequent appointments against the higher TTP that a shorter appointment interval can attain.

**Figure 3:**
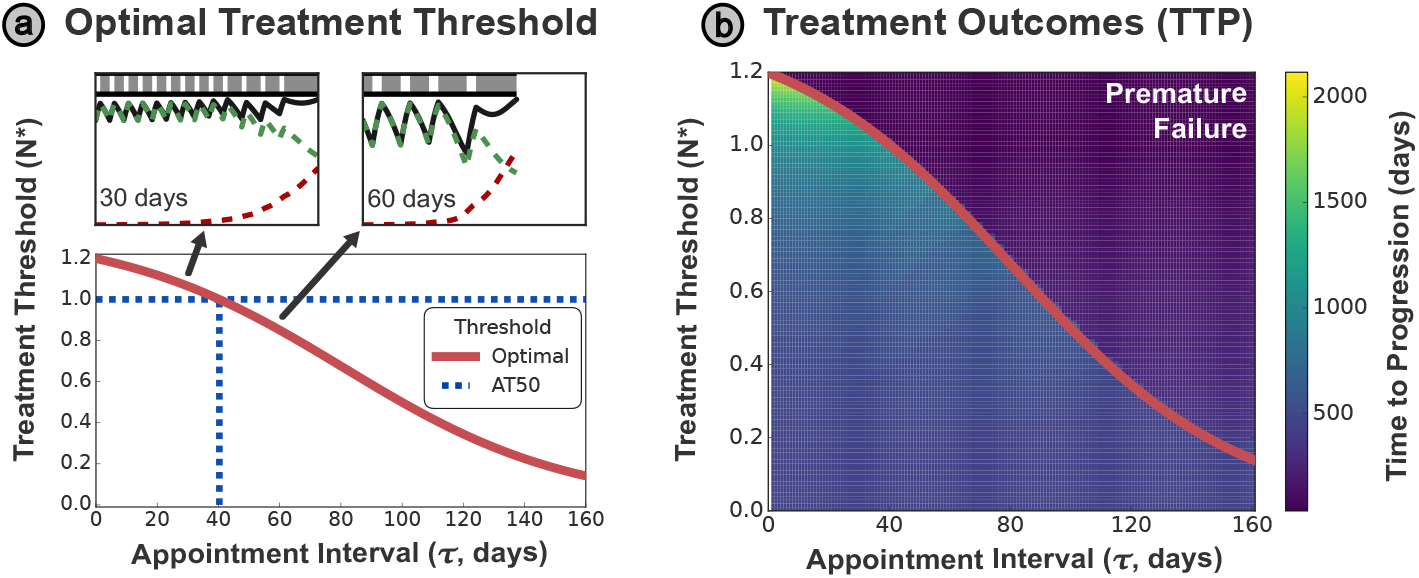
**(a)** The analytically derived optimal threshold *N*^*^ varies strongly with the appointment interval *τ*, meaning that AT50 is only optimal for a specific appointment interval (∼ 40 days). Intervals shorter than this benefit from a higher threshold tumor size, while those longer require a smaller tumor size. Examples of AT-N* are shown in the insets for *τ* = 30 and *τ* = 60 days - note that average tumor size is smaller under the 60 day interval, hastening tumor progression. **(b)** The curve *N*^*^(*τ*) can be plotted over the space of simulated TTP outcomes. To mitigate the sensitivity of the system on the first treatment timing, we simulate with an initial offset applied to the treatment schedule, and plot the minimum TTP observed across 20 simulations. The TTP is maximized at higher treatment thresholds, however, these are only possible for sufficiently frequent appointments. Treatment according to threshold values below *N*^*^ (for a given appointment spacing) will be sub-optimal, while thresholds above *N*^*^ have a risk of premature progression.

We systematically evaluate the TTP for different combinations of threshold size and appointment interval in Figure 3b. The occurrence of premature progression in such a grid search is highly sensitive to the timing of the discrete treatment intervals, and premature progression is dependent on when treatment re-evaluation occurs during the growth cycle (see Supplementary Figure S2). To obtain a more easily interpretable plot, we conduct multiple simulations of the TTP, adding a different offset period to the time points at which treatment is re-evaluated for each simulation. 20 different integer values (or *τ* values for *τ <* 20) for the offset duration, equally spaced between 0 and the appointment interval *τ*, are then used to simulate the TTP for each set of (*N*^*^, *τ*). See Supplementary Section S4 for a full motivation of this approach. We plot the minimum TTP across all 20 simulations, to replicate a ‘worst-case’ scenario for that particular treatment protocol (*N*^*^, *τ*).

Figure 3b reinforces the trend that a higher threshold tumor size results in a greater TTP. However, this only applies when the treatment plan is re-evaluated sufficiently frequently (i.e. when the time interval between appointments is below the optimal treatment curve defined by (7) and plotted in red). Insufficiently frequent clinical appointments for a given threshold tumor size are inherently risky, resulting in poorer treatment outcomes (driven by premature progression) observed in the region above the optimal treatment curve.

### 3.2 Waning Competition Model

For the more complex Waning Competition model introduced in Section 2.2, we see that the exponent of the competition term *α* affects the dynamics significantly. We illustrate this through simulation of AT-N* treatment protocols (with *N*^*^ = 0.7, *τ* = 60 days) for two different values of *α* in Figure 4a.

**Figure 4:**
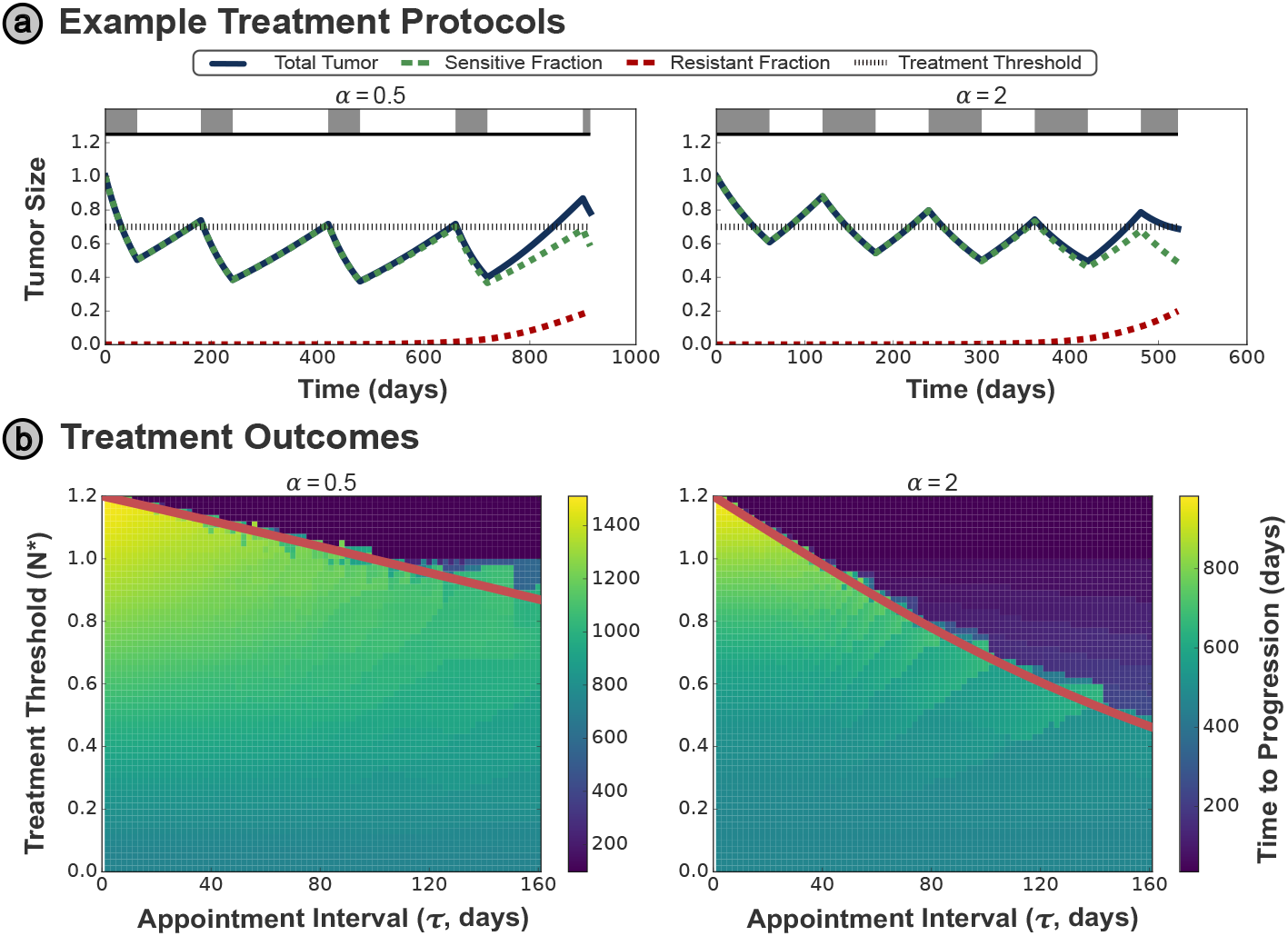
**(a)** Example treatment protocol (with *N*^*^ = 0.7, *τ* = 60 days) for the Waning Competition model under two different values of the critical exponent *α* — note that progression is defined by the growth of the resistant population to *R*(*t*) ≥ 0.1*K*_*R*_ for this model. **(b)** Treatment outcomes for differing *α* values, where the derived optimal threshold again accurately differentiates between premature progression above the optimal *N*^*^ curve (red solid line), and the sub-optimal TTP outcomes below the curve.

As outlined in Section 2.2, progression for this model is defined by Lu et al. (2024) as the growth of the resistant population to *R*(*t*) ≥ 0.1*K*_*R*_. However, it is also necessary to set an upper limit *N* (*t*) *<* 1.2*N*_0_ on the allowable total tumor size, because without this constraint it would be optimal to allow arbitrarily large drug-sensitive populations to suppress the growth of the resistant population.

In Supplementary Section S3.2, we derive the following relationship for *N*^*^ for this model:

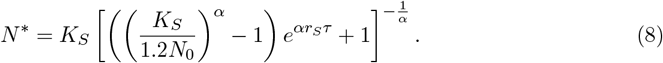

As previously discussed, this model also reduces to the generalized logistic model in the case where *γ* = 0. The threshold *N*^*^ considers the fastest possible growth for a tumor of given size but unknown composition, and this is maximized for *γ* = 0. Therefore, the expression for *N*^*^ given in (8) has no dependence on *γ*, and this expression also holds in the widely-used generalized logistic model.

This equation for *N*^*^ is overlaid on the treatment outcome space for multiple values of *α* in Figure 4b, and separates the regions with and without the possibility of premature progression. While this more complex tumor model can account for a wider range of dynamics than the Lotka–Volterra model considered in Section 3.1, the TTP space depicts the same trends in the *N*^*^ shown in the figure, and the optimal threshold framework outlined in Section 2.5 is equally applicable to this model. This illustrates the translatability of the optimal threshold *N*^*^, and the framework we propose for deriving this threshold, to different modeling assumptions and frameworks.

### 3.3 Stem Cell Model

The mathematical models examined so far assume that the treatment dynamics is primarily driven by expansion and contraction of two tumor subpopulation with differential, but constant, fitness in the presence and absence of drug. One implication of this assumption is that the rate of tumor re-growth off-treatment remains constant over time. However, a subset of patients in the AT clinical trial by Zhang et al. (2022) (from which the data in Figure 1b were also taken) display qualitatively different tumor dynamics that cannot be replicated mathematically by the previous tumor models, most notably an increasing regrowth rate off-treatment over time.

Exemplified for two patients in Figure 5a, we see that subsequent holiday periods are shorter, as the tumor size recovers at an increasing rate on subsequent cycles of AT, in contrast to the approximately-constant holiday lengths for the patients in Figure 1b. One plausible explanation for such behavior is provided by the Stem-Cell model proposed by Brady-Nicholls and Enderling (2022) (detailed in Section 2.2). This model treats tumor growth as driven by a separate stem cell population, instead of a drug-resistant sub-clone within the bulk of the tumor. While these stem cells are immune to the drug, they also drive the growth of the drug-sensitive, differentiated cell population directly. As stem cells accumulate over the simulation, the growth rate of the differentiated cells (that make up the bulk of the tumor) increases accordingly. Ultimately, the growth rate of the differentiated population (as stimulated by the stem cell population) exceeds the rate at which differentiated cells are killed by the drug, and the tumor grows under treatment to reach progression. Crucially, the principles of AT still apply to this model - progression is fundamentally driven (albeit indirectly in this model) by growth in the drug-resistant (stem cell) population, which may be suppressed by a larger population (close in size to the progression limit) of drug-sensitive (differentiated) cells.

**Figure 5:**
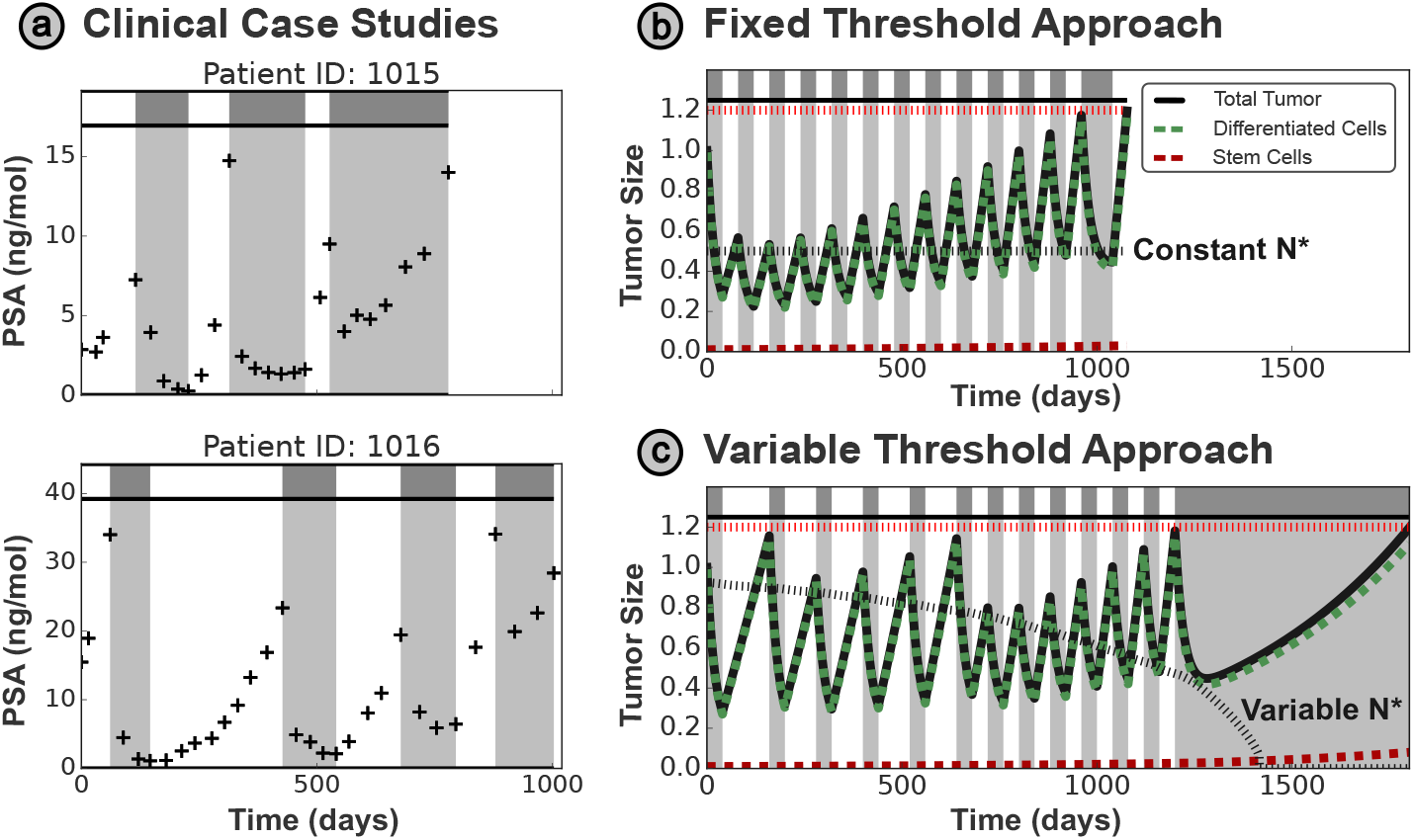
**(a)** Exemplar patient records from Zhang et al. (2022) demonstrate increasing rates of tumor rebound off-drug as treatment progresses. **(b)** This trend may be captured by the Stem Cell model of Brady-Nicholls and Enderling (2022), however, this behavior causes fixed threshold approaches to fail prematurely. In this plot, a fixed threshold of *N*^*^ = 0.6 for an interval *τ* = 40 days is suboptimal initially, with the total tumor size less than half of the progression limit. Despite this, the fixed threshold protocol still allows premature progression (i.e. tumor progression off-treatment) by the end of the simulation. **(c)** For this model, a time-varying treatment threshold is required to ensure maximal suppression of the resistant population without resulting in premature progression - this approach significantly increases TTP over fixed threshold approaches.

However, a fixed threshold approach is no longer appropriate for this model. In Figure 5b, we see that the threshold *N*^*^ = 0.5 is suboptimal for a treatment interval of *τ* = 40 days at the start of the simulation (maintaining a total tumor size significantly below the progression limit and hence weakening the competitive suppression of the drug-resistant stem cell population), while still failing to prevent premature progression occurring after approximately 1077 days. To address this issue, we extend our approach from Section 2.5 to allow for a threshold that changes over time to account for the change in tumor growth rate. The time-varying optimal threshold *N*^*^, as derived in Supplementary Section S3.3, is given implicitly by:

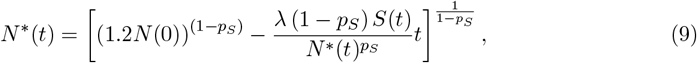

which we may solve numerically for *N*^*^(*t*).

While we cannot plot the TTP outcome over (*τ, N*^*^) space for time-varying *N*^*^(*t*), an exemplar treatment schedule for *τ* = 40 days is given in Figure 5c, which extends the TTP compared to the fixed threshold approach by 762 days. The concept of a time-varying treatment threshold is novel in AT, and was not required in an optimal treatment protocol for the previous models as they predict a constant rate of tumor regrowth over time. However, we have systematically shown that a time-varying threshold is optimal in certain modeling scenarios, such as the stem cell driven tumor growth in this model, and is necessary to account for specific patient dynamics observed in previous clinical trials of AT. These dynamics are not unique to prostate cancer, and have also been observed in melanoma, where mathematical models with phenotypic plasticity (switching between drug-sensitive and -resistant states) have been proposed to account for this behavior (Kim et al., 2021). This also raises wider questions regarding model selection for a given disease context, and whether multiple models should be used to capture qualitative heterogeneity in patient dynamics within a single study. Fundamentally, these results reflect the importance of tailoring mathematical models to observed experimental/clinical behavior of the target system, to enable the translation of analytic results such as optimal treatment protocols.

## 4 Discussion

With multiple ongoing clinical trials of AT (in skin (NCT05651828 - BCC Trial), prostate (NCT05393791 - ANZADPT Trial), and ovarian (NCT05080556 - ACTOv trial) cancers), it is of significant clinical interest to identify and characterize an optimal scheduling protocol for AT. However, previous work (Hansen et al., 2017; Viossat and Noble, 2021) to identify optimal AT approaches fails to account for discrete appointment intervals, which we show drastically modifies the ideal drug schedules.

We found that there is a trade-off between the appointment interval *τ* and the maximum attainable TTP, and introduce a threshold-based AT protocol, that maximizes the TTP for a given appointment interval. This threshold *N*^*^ is depends on patient-specific tumor parameters, motivating the clinical need for personalized AT frameworks that can adapt to the dynamics of each patient. We also show that different mathematical models may also be needed to capture qualitatively different patient dynamics, which can affect the optimal treatment strategy. To exemplify this, we identify patients with qualitatively different tumor dynamics (increases in the rate of tumor regrowth over time), and utilize a modeling framework that accounts for this behavior to propose a modified AT protocol where the treatment threshold may also vary over time. Our proposal that the best strategy may not just vary between patients, but also vary for a single patient throughout their treatment, is novel to AT; but we hope this may inspire other, innovative, approaches to drug scheduling in different disease contexts.

To implement personalized AT protocols clinically, we also need an approach to estimate the individual’s tumor parameters before treatment. In previous work (Gallagher et al., 2024), we have proposed a probing cycle to resolve this - all patients first undergo a standardized cycle of AT50, during which regular measurements of the tumor burden are taken. We may then fit our given mathematical model to these data to estimate the optimal *N*^*^ threshold for that patient, and these fits may be iteratively refined as more data are collected from subsequent treatment cycles.

Finally, we have discussed the role of modeling assumptions in this paper, and how that may affect conclusions on the optimal treatment schedule. Our finding that different tumor dynamics necessitate different AT scheduling approaches motivate the development of a systematic approach to identify the most suitable tumor model for a specific patient’s dynamics - bearing in mind that these dynamics may qualitatively vary between tumors in the same disease context undergoing the same treatment. One approach to this could be an established suite of complementary mathematical models that may be fit to the same dataset, allowing systematic comparison of predictions between each model (Strobl et al., 2023) — such an approach may be implemented through an ‘Evolutionary Tumor Board’ that directly interfaces between mathematicians and clinicians to recommend optimal treatment protocols personalized to specific patients under the clinicians’ care (Robertson-Tessi et al., 2023).

In summary, this paper illustrates the importance of accounting for clinical reality in the mathematical derivation of optimal treatment protocols, such as discrete patient monitoring. We show the importance of considering different underlying model assumptions based on the clinical data available, and demonstrate how these can lead to drastically different optimal treatment protocols, introducing a novel concept of an AT threshold that varies throughout a patient’s treatment. We hope this paper highlights the applicability of mathematical approaches to treatment scheduling in oncology, and inspires future work to translate these analytic approaches into clinically actionable protocols across a range of disease settings.

## Data Availability

All data presented in the present study are taken from the publicly accessible archive: https://doi.org/10.7554/eLife.76284, with a copy archived at https://archive.softwareheritage.org/browse/revision/ffa835ce8f4252d92a8c97f0e7324a1b6f87727b/?origin_url=https://github.com/cunninghamjj/Evolution-based-mathematical-models-significantly-prolong-response-to-Abiraterone-in-mCRPC&snapshot=25d81bfa294ef40b3eb90113cab9d43b90d4c91b
All code is available at https://github.com/KCGallagher/AT_Model_Comparison

https://archive.softwareheritage.org/browse/revision/ffa835ce8f4252d92a8c97f0e7324a1b6f87727b/?origin_url=https://github.com/cunninghamjj/Evolution-based-mathematical-models-significantly-prolong-response-to-Abiraterone-in-mCRPC&snapshot=25d81bfa294ef40b3eb90113cab9d43b90d4c91b

## Data Availability

All code is available at https://github.com/KCGallagher/AT_Model_Comparison

