## Supplemental Information for "Deriving Optimal Treatment Timing for Adaptive Therapy: Matching the Model to the Tumor Dynamics"

Oct 2024

#### S1 Model Parameters

##### S1.1 Lotka– Volterra Model

We adopt parameters from Strobl et al. (2021), which the authors showed generate simulations that are consistent with the response dynamics observed in metastatic prostate cancer patients treated with hormone therapy. In cases where a range was given, we have considered the parameters that correspond to a 'worst case' scenario when the tumor is least responsive to treatment. As characterized in Strobl et al. (2021), this occurs when there is no cost to resistance (i.e. $r_S = r_R$) and no cell turnover (i.e. $d_S = d_R = 0$). The full list of parameter values used are given in Table S1.

| Name | Description | Value/Range | Reference |
|---|---|---|---|
| $r_S$ | Sensitive cell proliferation rate | $0.027\,day^{-1}$ | Adopted from Zhang et al. (2017) |
| $r_R$ | Resistant cell proliferation rate | $1.0r_S$ | Zero cost scenario |
| $d_S, d_R$ | Natural cell death rate | 0 | Zero turnover scenario |
| $d_D$ | Drug-induced cell killing | 1.5 | Adopted from West et al. (2019) |
| $K$ | Tumor carrying capacity | 1 | Normalized to unity |
| $N_0$ | Initial tumor cell density | 0.75 | Rescaled by $K$ (Prokopiou et al., 2015) |
| $R_0$ | Initial resistant cell fraction | $0.001N_0$ | Adopted from Grassberger et al. (2019) |

Table S1: Parameter values used for the Lotka– Volterra model, taken from Strobl et al. (2021).

##### S1.2 Waning Competition Model

Parameter values for this model were taken from Lu et al. (2024), based on values that the authors obtained via a wide literature review, and are given in Table S2.

##### S1.3 Stem Cell Model

Values for the model parameters were taken from Brady-Nicholls and Enderling (2022), which the authors obtained via fitting to clinical data. Parameter values from Patient 1014 were selected for visualization. However, values from Patients 1002, 1005, 1010 and 1018 were also used to verify that our approach is robust to variation in parameter values (results not shown, but the same qualitative trends were observed).

| Name | Description | Value |
|---|---|---|
| $r_S$ | Sensitive cell proliferation rate | $0.00715\,day^{-1}$ |
| $r_R$ | Resistant cell proliferation rate | $0.023\,day^{-1}$ |
| $K_S$ | Carrying capacity for sensitive cells | 1.0 |
| $K_R$ | Carrying capacity for resistant cells | 0.25 |
| $d_S$ | Drug-induced sensitive cell killing | 2 |
| $d_R$ | Drug-induced resistant cell killing | 0 |
| $S_0$ | Initial sensitive cell density | 0.5 |
| $R_0$ | Initial resistant cell density | $2.5 \times 10^{-5}$ |
| $\alpha$ | Growth scaling term | 1.0 |
| $\gamma$ | Relative competition | $0.0021\,day^{-1}$ |

Table S2: Parameter values used for the Waning Competition model, taken from Lu et al. (2024).

| Name | Description | Value |
|---|---|---|
| $\lambda$ | Stem-like cell proliferation rate | $0.69\,day^{-1}$ |
| $p_S$ | Symmetric division probability | 0.0425 |
| $\alpha$ | Drug-induced sensitive cell killing | $0.0478\,day^{-1}$ |
| $S_0$ | Initial stem-like cell population | 10 |
| $D_0$ | Initial differentiated cell population | 1000 |

Table S3: Parameter values used for the Stem Cell model, taken from Brady-Nicholls and Enderling (2022).

### S2 Formal Analysis of Lotka –Volterra Model

#### S2.1 Stationary States

Here for completeness, we present the standard analysis we carried out for the Lotka –Volterra model. We consider the states where $\frac{dS}{dt} = 0, \frac{dR}{dt} = 0$; for simplicity we will also let $\delta(t) = (1 - d_D D(t))$. While this may vary over time, we will typically assume a constant value, denoted by $\delta$.

The non-trivial nullclines of this system (assuming non-zero values for each variable) are:

$$R = K\left(1 - \frac{d_S}{r_S \delta}\right) - S; \qquad R = K\left(1 - \frac{d_R}{r_R}\right) - S. \tag{S1}$$

Writing $\eta = \left(1 - \frac{d_S}{r_S \delta}\right)$ and $\mu = \left(1 - \frac{d_R}{r_R}\right)$, we obtain the nullclines:

$$R = \eta K - S; \qquad R = \mu K - S. \tag{S2}$$

From this we may extract three stationary points:

$$(S, R) = (0, 0) \quad , \quad (S, R) = (0, \mu K) \quad , \quad (S, R) = (\eta K, 0); \tag{S3}$$

in addition to the co-existence state $S + R = \eta K = \mu K$, in the special case where $\eta = \mu$.

We can think of $\eta$ and $\mu$ as the effective fitness of the sensitive and resistant populations respectively. Extending this analogy, negative values of $\eta$ or $\mu$ imply that the corresponding population cannot stably exist even in isolation, and corresponds to a fundamentally unfit population (i.e. a population that would become extinct in isolation, in the absence of competition with the other species). In a biologically relevant parametrization, we would expect both cell populations to reach a steady state in a nutrient-limited monoculture, and hence both $\eta$ and $\mu$ should be positive. The only exception to this is where $\delta < 0$, i.e. the drug has a net-killing effect on the sensitive population, driving it to extinction even in the absence of competitive effects.

#### S2.2 Stability Analysis

The Jacobian takes the usual form:

$$J = \begin{pmatrix} \frac{\partial \dot{S}}{\partial S} & \frac{\partial \dot{S}}{\partial R} \\ \frac{\partial \dot{R}}{\partial S} & \frac{\partial \dot{R}}{\partial R} \end{pmatrix} = \begin{pmatrix} r_S \left(1 - \frac{2S+R}{K}\right) \delta - d_S & -r_S \delta \frac{S}{K} \\ -r_R \frac{R}{K} & r_R \left(1 - \frac{S+2R}{K}\right) - d_R \end{pmatrix} \tag{S4}$$

**Point I: $(S, R) = (0, 0)$** Evaluating the eigenvalues of this at the stationary states, we find:

$$\lambda = r_S \delta - d_S,\, r_R - d_R. \tag{S5}$$

This point is only linearly stable if both eigenvalues $\lambda$ are negative. This corresponds to negative $\eta$ and $\mu$, which we interpret as fundamentally unfit species; otherwise this state is always unstable. This prevents us from driving the system to extinction in this model - i.e. complete tumor elimination is impossible.

**Point II: $(S, R) = (0, \mu K)$** From the Jacobian, we obtain the following eigenvalues:

$$\lambda = d_R - r_R,\, r_S \delta \frac{d_R}{r_R} - d_S. \tag{S6}$$

The first eigenvalue is always negative (provided $\mu > 0$), while the second eigenvalue is only negative when $\eta < \mu$. Therefore this resistant-only state is only stable when the effective fitness of the resistant population is greater than that of the sensitive population.

**Point III $(S, R) = (\eta K, 0)$** In this case, the Jacobian gives the following eigenvalues:

$$\lambda = d_S - r_S \delta,\, r_R \frac{d_S}{r_S \delta} - d_R \tag{S7}$$

The first eigenvalue is always negative (provided that $\eta > 0$), while the second eigenvalue is only negative when $\eta > \mu$. In contrast to Point II, the sensitive-only state is only stable when the effective fitness of the sensitive population is greater than that of the resistant population.

**Point IV: $S + R = \eta K = \mu K$** In this case, the complicated form of the Jacobian makes it difficult to easily gain insights. However, if we focus on the zero-turnover case (wherein $S + R = K$), we obtain interpretable eigenvalues:

$$\lambda = 0, -\frac{r_R R + \delta r_S S}{R + S} \tag{S8}$$

This second value is always negative (provided $\delta < 0$), while the zero eigenvalue (with corresponding eigenvector $(-1, 1)$) allows perturbations within this state; that is, the steady states are neutrally stable and perturbations to this state move the system to another steady state along this continuum. We find numerically (not shown) that this trend is replicated in the case of non-zero turnover, provided that $\eta = \mu$.

#### S2.3 Parameter Regimes

We now summarize the results above. It is worth highlighting that the stability of each state depends on $\delta$, which we take to be constant in the analysis below, and modification of the drug dose could alter the stability of a given state.

**Case I $\eta > \mu$** In this case, the only stable stationary state is the sensitive-exclusionary state $(S, R) = (\eta K, 0)$. This parameter regime corresponds to $\frac{r_R}{d_R} < \frac{d_S}{r_S}\delta$ - i.e. resistant cells have higher turnover (than the drug limited 'effective' turnover of the sensitive cells). For equal (non-zero) natural death rates, this case corresponds to a fitness advantage for sensitive cells - i.e. $r_S\delta > r_R$. This fitness advantage is unlikely to occur under drug treatment (unless sensitive cells are sufficiently fit to out-compete resistant cells when sensitive growth is limited by drug), and cannot occur if $d_D \geq 1$ - i.e. drug has a net killing effect - as $\delta < 0$.

**Case II $\eta < \mu$** In this case, only the resistant-exclusionary state $(S, R) = (0, \mu K)$ is stable. This parameter regime corresponds to $\frac{d_S}{r_S\delta} > \frac{d_R}{r_R}$ - i.e. resistant cells have lower turnover than the drug limited 'effective' turnover. For equal (non-zero) natural death rates, this case corresponds to an effective fitness advantage for resistant cells under drug treatment - i.e. $r_R > r_S\delta$. This fitness advantage is guaranteed if the drug has a net killing effect ($d_D > 1$) - as $\delta < 0$, but occurs generally when the impact of the drug outweighs the baseline proliferation advantage of sensitive cells.

**Case III $\eta = \mu$** This parameter regime results in a continuum of steady states given by $S + R = \eta K = \mu K$, defining a co-existence state that only exists for the direct equality $\eta = \mu$. This special case corresponds to $\frac{d_S}{r_S\delta} = \frac{d_R}{r_R}$. Primarily, this occurs if there is no cellular turnover ($d_R = d_S = 0$), or for a zero-cost case off-treatment, where $r_R = r_S$ (in which case both cell populations are identical). However, for drugs that reduce the net growth rate of sensitive cells rather than having a net killing effect ($d_D < 1$), there also exist sets of parameters that satisfy this condition for non-zero cost and turnover.

This state is structurally unstable, as any uncertainty or variation in the parameter values will perturb the system into a different state. For this reason, we will neglect Case III when considering clinically-implementable treatment strategies.

##### S2.4 Overall Dynamics

These three cases correspond to the three pairs of nullclines plotted in Figure S1a. We can formally consider the transition between these states as a bifurcation, where the number and nature of the stationary states changes as $\eta - \mu$ changes sign. Varying these two parameters (representing the relative fitness of each population) relative to each other (plotted in Figure S1b) displays this change in stability of those states. Each species has a stable non-zero stationary state when its relative fitness is greater than that of the other species, while coexistence is possible when the species have the same relative fitness.

In all practical implementations of adaptive therapy, the progression limit $1.2N_0$ will be less than $min(\eta K, \mu K)$, and so the tumor will continue to grow to this progression limit for any treatment schedule. Adaptive therapy instead aims to drive the tumor over to the upper right region of phase space (Figure S1c) where the net tumor growth rate ($\frac{dN}{dt}$) on treatment is reduced, to delay progression for as long as possible.

The tumor growth is controlled via modulation of the drug term $\delta$: AT switches between Cases I and II to keep $(1.2N_0 - N(t))$ small but strictly positive (i.e. a tumor that remains marginally smaller than the progression limit). To do this, we will implement an optimal tumor threshold - when the tumor is larger than this threshold size, the drug dose should be sufficient to ensure that the system is in Case II, where the overall size of a primarily-sensitive tumor will shrink. Below the threshold size, the drug dose should be lowered (or removed entirely in the case of binary dosing protocols) to return to Case I dynamics, which corresponds to resensitization of the tumor. In Section S3.1, we will derive this optimal size for the Lotka – Volterra model.

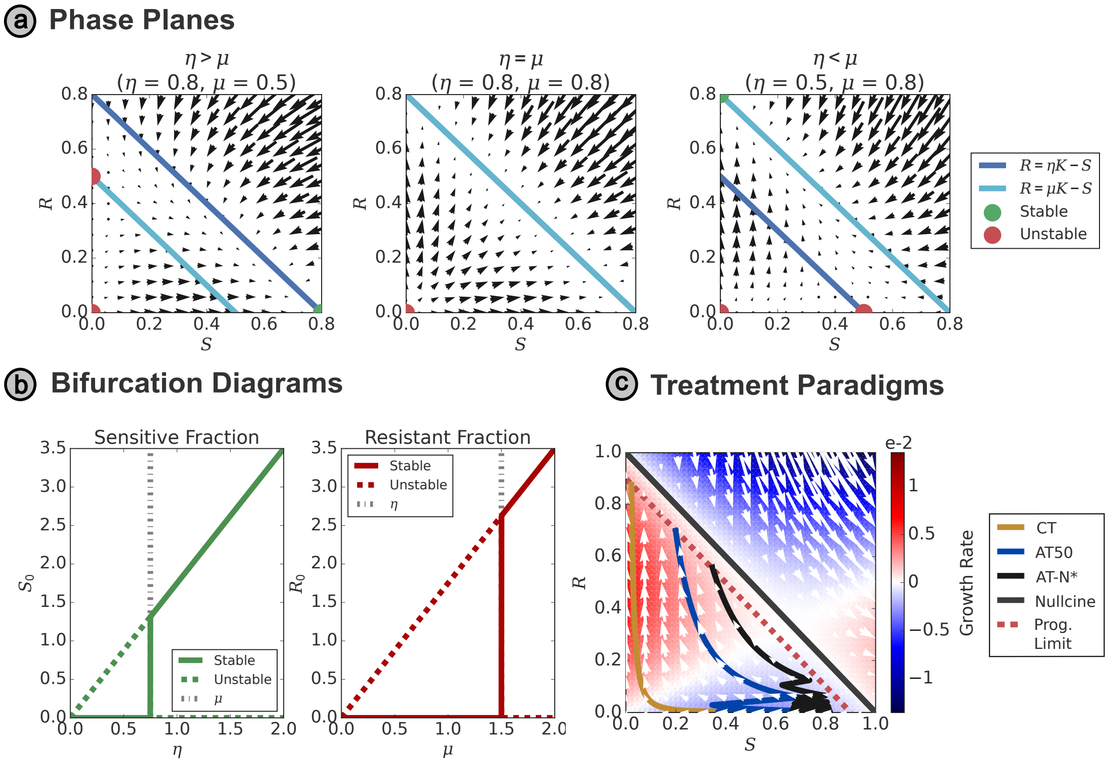


Figure S1: **(a)** Phase plane plots for each of the three cases described in Section S2.3, with stationary states depicted by circles in each one. In the $\eta = \mu$ case, the two nullclines overlap, resulting in a continuum of stationary states. **(b)** Each species has a stable non-zero stationary state, when its 'relative fitness' is greater than that of the other species. Coexistence is possible when the species have the same 'relative fitness', while the other species has a stable non-zero stationary state (of constant size) below the bifurcation. **(c)** A comparison of different treatment protocols in phase space. AT maintains the tumor state close to the progression limit, where the net growth rate of the tumor on treatment is lower, resulting in delayed progression.

### S3 Optimal Threshold Derivation

#### S3.1 Lotka – Volterra Model

We may estimate the critical treatment interval $\tau^*$, assuming $R = 0$ such that the total tumor size $N$ is equal to the sensitive population $S$ (we will justify this assumption in Section S3.1.1). Note that in this (and all subsequent) analysis, we also assume that the total tumor size is less than the carrying capacity ($N(t) < K \, \forall \, t$) - a reasonable assumption given the clinical context of this work that assumes the tumor will tend to naturally grow when no drug is present.

Rewriting (1) with $S \approx N$ and $R \approx 0$, we obtain:

$$\frac{dN}{dt} = r_S N \left(1 - \frac{N}{K}\right) - d_S N. \tag{S9}$$

Integrating over the critical treatment period $\tau^*$:

$$\int_{N^*}^{1.2N_0} \frac{1}{r_S N \left(1 - \frac{N}{K}\right) - d_S N} dN = \int_0^{\tau^*} dt = \tau^*,$$

which may be evaluated to obtain:

$$\tau^* = \frac{1}{r_S - d_S} \, ln\left[\frac{1.2N_0}{N^*}\frac{K(r_S - d_S) - r_S N^*}{K(r_S - d_S) - 1.2 r_S N_0}\right]. \tag{S10}$$

In practice, the interval between appointments is often determined by clinical availability and practical restrictions, so it is much more practical to personalize the threshold tumor size within the AT protocol to each patient. We therefore rearrange (S10) to obtain an expression for the optimal treatment threshold based on a given appointment interval $\tau$:

$$N^* = \frac{K(r_S - d_S)}{\left(\frac{K(r_S - d_S)}{1.2N_0} - r_S\right)e^{(r_S - d_S)\tau} + r_S}. \tag{7 revisited}$$

##### S3.1.1 Non-Negligible Resistant Fractions

In Section S3.1, we have assumed the resistant cell population ($R$) is negligible. While this may be true initially, this subpopulation will ultimately grow sufficiently to result in relapse. We therefore show that the critical appointment interval $\tau$, given by (S10) still holds for any tumor composition $N = S + R$.

Let us suppose $r_S = r_R + \epsilon; d_R = d_S + \zeta$, where $\epsilon \geq 0; \zeta \geq 0$ - i.e. both the growth rate $r_S$ and the death rate $d_S$ of the sensitive cells are greater than or equal to the corresponding values for the resistant cells. This gives the following expression for the total tumor size $N$:

$$\frac{d}{dt}(S + R) = \left(1 - \frac{N}{K}\right)[r_S S + r_S R - \epsilon R] - d_S S - (d_S + \zeta) R. \tag{S11}$$

As before, we constrain this analysis to $N < K$, i.e. for tumor sizes less than the carrying capacity, because $N = K$ corresponds to non-positive growth rates of both the sensitive and resistant populations. We can therefore rewrite (S11) as:

$$\frac{d}{dt}(S + R) = r_S N\left(1 - \frac{N}{K}\right) - d_S N - R\underbrace{\left[\epsilon\left(1 - \frac{N}{K}\right) + \zeta\right]}_{+ve}, \tag{S12}$$

where the final term is non-negative. Contrasting to the growth rate of a fully sensitive tumor (S9), we can see that the off-treatment growth rate of the mixed tumor is necessarily less than or equal to that of the fully sensitive tumor. This corresponds to a decrease in the critical appointment interval (S10), since the mixed tumor will grow slower than the fully sensitive tumor and so we will have more opportunities to intervene, and hence an increase in the maximum allowable treatment threshold (7). The critical threshold given by (7) is therefore exact when the resistant population is negligible (or there is no fitness difference between the two populations) and an overestimate otherwise. Given the resistant fraction of the tumor will change over time, and is often not known at the start of treatment, we therefore conclude that (7) represents a 'worst-case' scenario.

#### S3.2 Waning Competition Model

As the progression threshold in this model is given by $R(t) \geq 0.1K_R$, we may apply the same procedure to derive $N^*$ as for the Lotka – Volterra model. We also retain the constraint that the total population $N(t) := S(t) + R(t)$ must be less than the conventional progression threshold $1.2N_0$ - while this was not specified by Lu et al. (2024), it is necessary to ensure tolerable tumor sizes. Without an upper bound on the allowable size of the tumor, the optimal strategy would be to maximize competition by simply leaving the tumor untreated at all times so that the sensitive population may grow unchecked. We therefore maximize the sensitive population subject to this constraint, to reduce $\frac{dR}{dt}$, and hence the time to progression.

Given that $r_S > r_R$, we may apply the same logic as in Section S3.1 and consider the growth rate of a wholly sensitive tumor (with negligible resistant fraction) and minimal inter-species competition ($\gamma = 0$), for the limiting case of fastest tumor recovery (and highest risk of premature failure). In this case, the growth dynamics simply reduce to:

$$\frac{dS}{dt} = r_S S \left[1 - \left(\frac{S}{K_S}\right)^\alpha\right],$$

which gives the integral (for the non-dimensionalized size $\hat{S} = \frac{S}{K_S}$):

$$\tau = \frac{1}{r_S} \int_{\frac{N^*}{K_S}}^{\frac{1.2N_0}{K_S}} \frac{1}{\hat{S}\left(1 - \hat{S}^\alpha\right)} d\hat{S}.$$

Integrating, we obtain:

$$N^* = K_S \left[\left(\left(\frac{K_S}{1.2N_0}\right)^\alpha - 1\right) e^{\alpha r_S \tau} + 1\right]^{-\frac{1}{\alpha}}. \qquad \text{(8 revisited)}$$

Lu et al. (2024) only consider the case in which $\alpha = 1$; in this case (8) reduces to:

$$N^* = \frac{K_S}{\left(\frac{K}{1.2N_0} - 1\right) e^{r_S \tau} + 1}.$$

##### S3.3 Stem Cell Model

While in previous computations we have assumed that the drug-resistant population (i.e. the stem cell population $S(t)$) is negligible, that is not possible for this model as the growth rate of the differentiated cells is directly proportional to the stem cell population.

We instead rewrite (3) during periods without treatment solely in terms of $N, S$:

$$\frac{dN}{dt} = \frac{dS}{dt} + \frac{dD}{dt} = \lambda S,$$
$$\frac{dS}{dt} = \left(\frac{S^2}{N}\right) p_S \lambda.$$

Hence, we have the separable equation:

$$\frac{dN}{dS} = \frac{N}{p_S S}, \qquad \text{(S13)}$$

from which we obtain:

$$N(t)^{p_S} = \frac{N(0)^{p_S}}{S(0)} S(t). \qquad \text{(S14)}$$

Using this, we may derive an exact solution for the drug-free growth of the whole tumor:

$$N(t) = \left[N(0)^{(1-p_S)} + \frac{\lambda\left(1 - p_S\right) S(0)}{N(0)^{p_S}} t\right]^{\frac{1}{1-p_S}}.$$

In particular, we may consider the drug-free growth after some time $t$, starting from a tumor size $N^*$. As the tumor size must reach the progression limit $1.2N(0)$ in the appointment interval $\tau$ to trigger premature progression, we obtain the time-dependent expression for $N^*$:

$$N^*(t) = \left[(1.2N(0))^{(1-p_S)} - \frac{\lambda\left(1 - p_S\right) S(t)}{N^*(t)^{p_S}} \tau\right]^{\frac{1}{1-p_S}}, \qquad \text{(9 revisited)}$$

which we may solve numerically for $N^*(t)$. While the time dependence for $N^*$ is contained within $S(t)$, it would also be possible to extract this from (S13), provided we integrate over the treatment history of the patient.

#### S4 Varied Offset Simulations

When the treatment threshold used for a model is larger than the corresponding optimal threshold derived in Section S3, there is a risk of premature progression. However, this is not guaranteed, as our derivation for $N^*$ assumes a 'worst-case' scenario of the tumor size being just below $N^*$ at the start of a treatment interval. Under threshold-based AT with discrete intervals between appointments, it is equally possible for the tumor size to undershoot the threshold while on treatment as it is to overshoot the threshold while off treatment, and in many cases these effects will cancel out.

To demonstrate that this effect results in extreme sensitivity to the timing of treatment re-evaluation, we may also include an offset period (of duration $t < \tau$) at the start of the simulation - this merely changes the dates upon which treatment is re-evaluated and does not affect the optimal threshold. For example, an offset of 20 days (when $\tau = 60$ days) means that treatment still starts at $t = 0$ but is subsequently re-evaluated at time points $t = 20, 80, 140...$ days). The regular threshold-based AT treatment protocol will be applied in the first treatment period, such that treatment will be given for the offset period if the initial tumor size $N(t = 0)$ is greater than the threshold size). Applying an offset period is equivalent to changing the initial size and composition of the tumor, while leaving the progression threshold and tumor dynamics unchanged.

An example of this is given in Figure S2a. Here we consider simulations of the Lotka–Volterra model with an appointment interval of 60 days, where the corresponding optimal threshold is $N^* = 0.85$. To illustrate the risk of premature progression when a threshold greater than the optimal threshold $N^*$ is used, we simulate threshold-based AT with a slightly greater threshold of $N = 0.87$. We see that the duration of the offset period drastically impacts the overall TTP - with an offset of 20 days the treatment schedule maintains control of the tumor until natural progression (driven by the resistant population), whereas differing offset values can result in premature progression at different time points, with an offset of 40 days approximately halving the maximal TTP. This extreme variation in TTP is also apparent in the full treatment outcomes space; when considering all combinations of $(\tau, N^*)$ in Figure S2b we see that the occurrence of premature progression for thresholds greater than the optimal threshold is unpredictable. To mitigate this noise, we simulate each treatment schedule (defined by $\tau, N^*$) with multiple different offset values, and plot the mean TTP in Figure S2c. This figure displays a much smoother transition in TTP across the optimal threshold boundary.

Our derivation of the optimal threshold does not strictly predict the combinations of $(\tau, N^*)$ where premature progression definitely will occur, rather it specifically allows us to differentiate between treatment schedules where it cannot occur and schedules where there is a possibility of premature progression occurring. This is intended to provide a minimal-risk treatment schedule, where there is no risk of premature progression even if clinical appointments are rescheduled, provided the maximal gap between appointments is not longer than $\tau$. For this reason, we present the treatment outcome spaces in the main text based on the minimum TTP obtained across a range of offset values, to replicate the 'worst-case' scenario in the clinic. In each case, we simulate the TTP subject to 20 different offset periods, of integer durations equally spaced between 0 and $\tau$, and plot the minimum TTP attained. If the appointment interval $\tau$ is less than 20 days, then we instead simulate for an offset given by each integer between 0 and $\tau - 1$ inclusive.

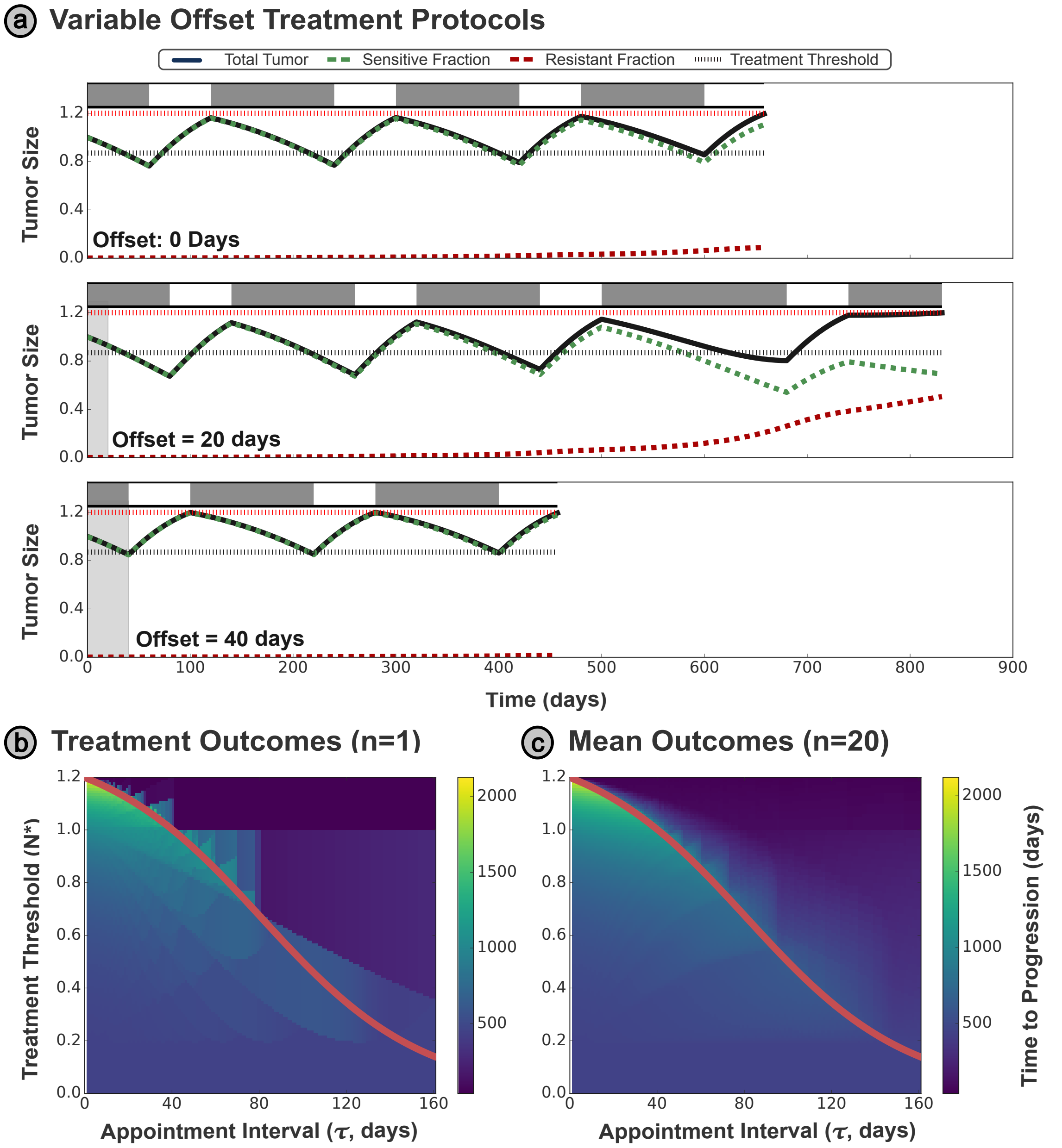


Figure S2: **(a)** Example treatment protocols (with a threshold size $N = 0.87$, $\tau = 60$ days) for the Lotka–Volterra model — this threshold is chosen to be slightly greater than the optimal value $N^* = 0.85$. Including varied time offsets in the appointment schedule demonstrates the sensitivity of the system and TTP to the precise timing of treatment. **(b)** Treatment schedules with no time offset display highly variable TTP outcomes just above the optimal threshold. **(c)** Computing the mean TTP from multiple simulations with different time offsets at the start of treatment allows us to mitigate the sensitivity to the treatment timing.
